# An expanded high throughput RT-PCR assay to rapidly identify all known SARS-CoV-2 variants of concern using melting temperature coding

**DOI:** 10.1101/2022.01.18.22269424

**Authors:** Padmapriya P Banada, Raquel Green, Sukalyani Banik, Deanna Streck, Ibsen Montalvan, Robert Reiss, Robert Jones, Salvatore A. E. Marras, Soumitesh Chakravorty, David Alland

**Affiliations:** Public Health Research Institute, Rutgers New Jersey Medical School, Newark, NJ; Institute of Genomic Medicine, Rutgers New Jersey Medical School, Newark, NJ; University Hospital, Newark, NJ; Division of Infectious Diseases, Department of Medicine, Rutgers New Jersey Medical School, Newark, NJ; Craic Computing LLC, Snohomish, WA; Cepheid, Sunnyvale, CA

**Keywords:** SARS-CoV-2, Delta, Omicron, N501Y, E484K, E484A, L452R, melting temperature (Tm), screening test, surveillance

## Abstract

**Background:** The rapid emergence of new vaccine-resistant SARS-CoV-2 variants of concern (VOC) requires an equally rapid deployment of diagnostic tests to specifically identify each VOC as soon as it arises. Here, we report an expanded version of our previously described sloppy molecular beacon (SMB) Alpha/Beta/Gamma RT-PCR melting temperature (Tm) signature-based assay, which now includes modifications that allow specific detection of Delta (B.1.617.2) and Omicron (B.1.529) VOCs.

**Methods:** We developed a dual SMB assay (SMB-452), which targeted the T22917G (L452R) mutation in the SARS-CoV-2 spike protein to specifically detect the Delta VOC. We also identified a Tm profile in our existing SMB-501 and SMB-484 assays (which detect mutations in codons 501 and 484 of the SARS-CoV-2 spike protein, respectively) that differentiate the Omicron-specific N501Y (A23063T) and E484A (A23013C) mutations from both wild type (WT) and other VOCs. The entire six SMB three-codon assay was tested using reference SARS-CoV-2 RNAs. The assay was then validated using clinical samples from COVID-19 patients tested with a LightCycler 480 (LC480) (74 samples), Bio-Rad CFX96 (34 samples), Rotor-Gene Q (Qiagen) (34 samples) and an ABI-7500 (34 samples) RT-PCR instruments. Six SMB Tm results were then inputted into an Excel Analysis tool to generate specific VOC identifications.

**Results:** The limit of detection (LOD) for the new SMB-452 assay, which specifically identified the Delta variant was 1 genomic equivalent (GE) per reaction. The LODs of the SMB-501 and SMB-484 assays, which detect Omicron were 100 and 10^3^ GE respectively. Clinical validation of the 3-codon assay in the LC480 instrument showed the assay detected 94% of the samples as WT or VOCs in clinical samples and 6% of the tests producing indeterminate results. None of the samples were incorrectly identified as WT or as a different VOC. Thus, excluding samples with indeterminant results, the assay was 100% sensitive and 100% specific compared to sequencing. There was also 100% concordance between the LC480, BioRad, ABI and Qiagen results, excluding negative or indeterminate results; however, the Qiagen assay had significantly more indeterminates than the other assays.

**Conclusion:** This new assay can serve as a robust diagnostic tool for selecting appropriate monoclonal antibody therapy and rapid VOC surveillance.

## Introduction

The global emergence of SARS-CoV-2 variants of concern (VOC), B.1.1.7 (20I, Alpha), B.1.351 (20H, Beta), P.1 (20J, Gamma), B.1.617.2 (21A, Delta), and B.1.1.529 (21K/ Omicron) have been responsible for a series surges in reported COVID-19 cases (1). The Omicron variant, first identified as a VOC by the WHO in November 2021 (2) is now dominating the United States, accounting for 95% of the COVID-19 cases as of Jan 04, 2021 (3). Several studies have indicated that the Alpha, Beta, Gamma and Delta variants are more transmissible and possibly more virulent than previous SARS-CoV-2 strains (4-7), and the CDC and others have indicated that the Omicron variant may spread twice as rapidly as Delta, but published data are unavailable at the time of this manuscript preparation. The VOCs, and especially Omicron, may confer resistance to therapeutics and decreased vaccine efficacy (7-11) due to the presence of key mutations in the spike protein (12-16). Although the United States has recently increased it capacity to track variants by genome sequencing, only between 2% to 10% of COVID-19 cases are being sequenced (17, 18) and the results of these efforts are often delayed by up to 10 days (19), threatening their utility in preventing further spread or providing real-time therapeutic guidance. A more rapid approach to screen for potential Omicron variants takes advantage of specific RT-PCR COVID-19 tests, which fail to detect S-gene target sequences in Omicron variants. Samples that are negative for the spike target but positive for another SARS-CoV-2 sequence identify presumptive Omicron infections, which are then confirmed by DNA sequencing (1). However, this type of test only identifies Omicron variants, and it still requires sequence confirmation and the associated time delay.

We have previously demonstrated an RT-PCR based method that uses sloppy molecular beacons (SMBs) combined with melting temperature (Tm) code analysis to detect mutations in codons 501 and 484 of the SARS-CoV-2 spike protein. We showed that this assay could specifically identify WT SARS-CoV-2 and the Alpha, Beta and Gamma variants (20). Delta and Omicron have developed additional variant defining mutations. For Delta, these include the spike protein mutations E156-, F157-, L452R, T478K, D614G, P681R, D950N; for Omicron, these include the spike mutations H69-, V70-, K417N, T478K, E484A, N501Y, D614G, P681H. As our earlier assay only targeted mutations in codons 501 and 484 of the spike protein, it was unable to identify Delta variants. However, we expected that a new assay, which could detect mutations at codon 452 should be able to identify Delta variants, and that our existing codon 484 assay would be able to specifically identify Omicron variants if it was able to distinguish the WT 484 sequence present in both WT and Alpha strains from the E484K mutation present in Beta and Gamma, and the E484Q mutation present in Kappa from the E484A mutation present in Omicron (21). Our codon 501 assay would also be able to provide additional robustness by distinguishing Delta from Omicron since Omicron contains the N501Y mutation also present in Alpha, Beta, and Gamma while Delta does not (5, 12, 22, 23). Thus, we expected that a simple combination of assays targeting mutations at codons 501, 484, and 452 of the spike protein would be able to specifically identify all the current SARS-CoV-2 VOCs, providing a rapid diagnostic and epidemiological tool.

Here, we describe an expanded assay to identify and distinguish Delta and Omicron variants along with Alpha, Beta, and Gamma, with high specificity and sensitivity. We demonstrate that this approach is flexible and can be used for detecting VOCs using both SARS-CoV-2 RNA and clinical samples. We further confirmed assay performance in four different RT-PCR instruments commonly available in diagnostic laboratories. Adaptation of our assay will enable real-time detection of SARS-CoV-2 variant spread, without the need for whole genome sequencing on all samples.

## Methods

### Ethical considerations

The usage of de-identified clinical samples from RT-PCR confirmed COVID-19 positive and negative patients in this study was approved by the Rutgers University institutional Review Board under protocol numbers 20170001218 and 2020001541.

### Viral cultures and RNA

SARS-CoV-2 RNA or viral cultures (USA WA1/2020 (WT, NR52285), B.1.1.7 (Alpha, NR54000), B.1.351 (Beta, NR-55282), P.1 (Gamma, NR-54982) B.1.617.2 (Delta, NR-55611) and B.1.529 (Omicron, NR-56461) were obtained from BEI Resources, NIAID (Manassas, VA). RNA was isolated from the variant strains in a BSL3 laboratory, using RNAdvance viral RNA extraction kit (Beckman Coulter, Indianapolis, IN). The extracted RNA was quantified against a standard curve generated with the N gene-specific real time RT-PCR assay. RNA extracted from B.1.529 Omicron variants hCoV-19/USA/GA-EHC-2811C/2021 (GISAID: EPI_ISL_7171744) and hCoV-19/USA/WI-WSLH-221686/2021 (GISAID: EPI_ISL_7263803) were a kind gift by Dr. Vincent Munster, Rocky Mountain labs, National institutes of Health (NIH).

### Assay design, primers and probes

SMBs and primers verified to detect mutations in codon N501 and E484 were used as reported from our earlier publication (20). An additional assay to detect the L452R (T22917G) mutation present in B.1.617.2 (Delta) was designed similarly as described previously (20). Briefly, a total of 412,389 high quality SARS-CoV-2 genome sequences deposited in GISAID (24) as of Feb 19, 2021, were analyzed using BLAST (25) and aligned with MAFFT (26). Primers and probes were designed on the basis of sequence conservation using Primer3 program (27) to amplify a 122 bp region flanking the position 22917 (452 codon) in the reference strain (GenBank accession number MN908947). SMB probe design was performed using the web servers DNA mfold (http://www.unafold.org/mfold/applications/dna-folding-form.php) and DINAmelt (http://www.unafold.org/hybrid2.php) to predict the probe folding structures and probe-target hybrid Tm values respectively. Similar genome analysis was performed for the B.1.529 (Omicron) strain comparing it to other variant strains at the primer/probe binding regions. A total of 4048 high quality sequences were analyzed for 484 mutations and 3,964 high quality sequences for 501 mutations. All analyzed genomes for 452 were wildtype. In-silico two state melting hybridizations was performed to understand the Tm variations using DINAmelt application. The final list of primers and probes used in this study are listed in Table 1. Primers were obtained from Millipore Sigma and SMBs were synthesized by LGC Biosearch technologies (Petaluma, CA). An internal control (IC) assay developed by CDC (28, 29), targeting the human RNaseP gene was simultaneously performed for each extracted RNA specimen as a separate reaction in a separate well, using the *Taq*Man real-time PCR assay probe tagged with FAM at the 5’ end and Dabcyl quencher at the 3’ end.

**Table 1.**
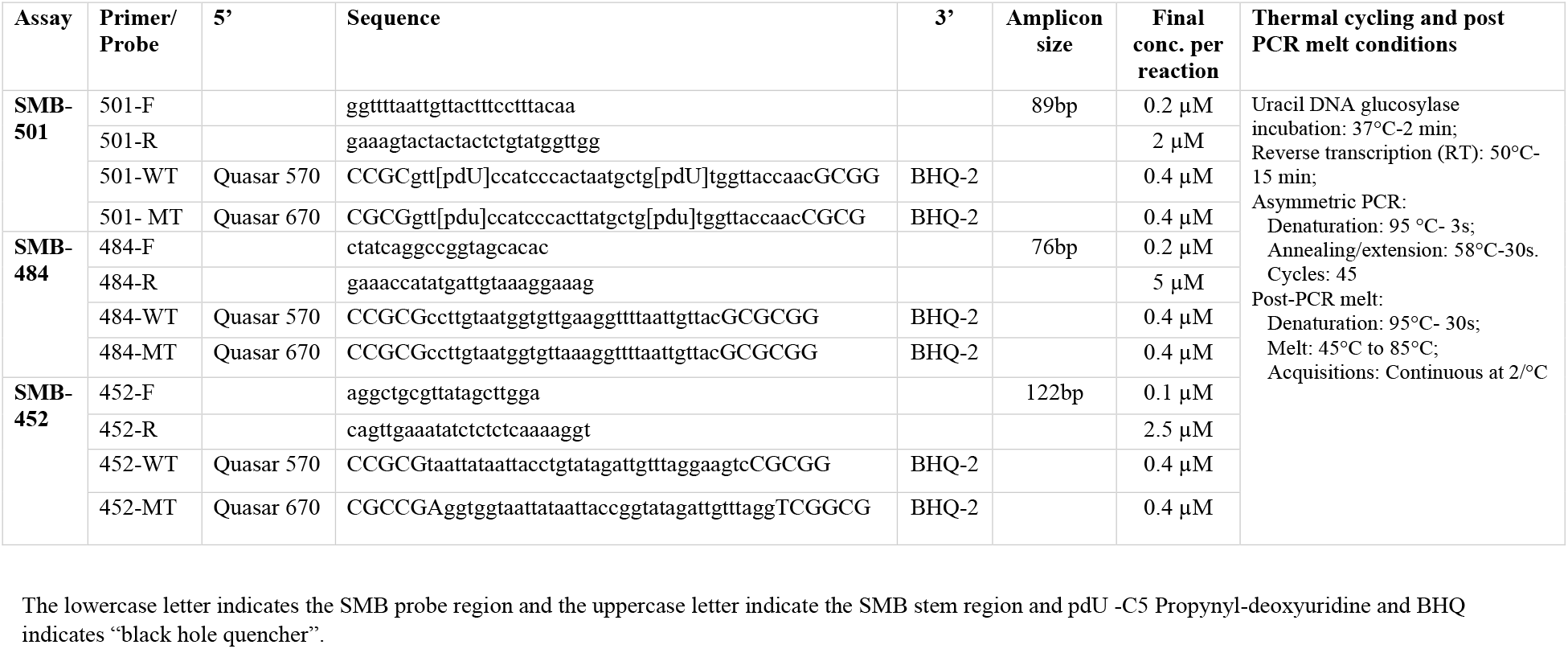
Primers, probe, and PCR conditions.

### SMB-assay formulation

TaqPath™ 1-Step RT-qPCR Master Mix, CG (ThermoFisher Scientific, Waltham, MA) was supplemented with the assay primers and probes at final concentrations as mentioned in Table 1 for an asymmetric one-step RT PCR. A 1 µl of the template RNA was added per 20 µl reaction.

### Analytical sensitivity

The pre-quantitated genomic RNA from the SARS-CoV-2 B.1.617.2 (Delta) and B.1.529 (Omicron) were diluted in Tris-EDTA (TE) buffer. For the background matrix, total nucleic acids were extracted from a SARS-CoV-2 negative nasopharyngeal (NP) specimen (confirmed negative by Xpert Xpress SARS-CoV-2 test). RNA from the Delta/Omicron were spiked to the negative matrix and diluted to achieve the final concentrations of 10^5^, 10^4^, 10^3^, 100, 10, 1, and 0.1 GE/µl. A 1µl aliquot of this mix was added to 19µl of the one-step RT-PCR mix containing the primers and probes and was evaluated in the SMB-501/SMB-484/ SMB-452 assays.

### Excel analyze tool for identification of SARS-CoV-2 variants

For identification and classification of the VOCs, we modified a Microsoft Excel based program originally published by Chakravorty et al. (30). Briefly, the program finds the closest match between the Tm signature from the patient samples to that of the reference VOCs. A distance index (D-value) is calculated based on the difference in values between the reference and the unknown. A D-value of <5 was considered in these studies as a perfect match and ≥5 was classified as “indeterminate”. The program uses the Ct value of the internal control (IC) to assess failed run and a successful run. An “invalid” call is made if the IC fails to generate a Ct along with the SMB probes and is called “negative” if all probes are negative except for the IC. For VOC classification, the Tm signature values that are generated from each of the six SMB probes (two SMB for each of the three codons) are entered and the tool generates an output result of either ‘SARS-CoV-2 negative’/’Wild type (Ancestral)’/’B.1.1.7 (Alpha)’/ ‘B. 1.351 (Beta)’/’B.1.617.2 (Delta)’/ or ‘B.1.529 (Omicron)’. An ‘indeterminate’ output is obtained if the Tm values are outside of the reference window or if a Tm value of zero is entered for >2 SMB probes due to the failure of these probes to generate a Tm. However, if only 1 or 2 of the SMB probes fail to generate a Tm, the tool matches the remaining Tm values to the closest reference and reports the identified VOC as ‘presumptive’. This program file could be shared upon request to the readers.

### Assay protocol

All assays were run as separate reactions as a 4-well test including the internal control. Each test sample was run in replicates of 4 in 384-well plates in a Roche LightCycler 480 (Roche, Indianapolis, IN). The one-step RT-PCR amplification was performed with same PCR conditions described previously (20), and mentioned in Table 1. The total assay time was 1h 17min. Tm values obtained from the instrument for each SMB-assay from both WT and MT probes, were exported and identified using the Excel analyze tool.

### Clinical specimen evaluation and RT-PCR instrument feasibility

A total of 74 specimens containing deidentified nasopharyngeal (NP) swabs, nasal swabs, and saliva obtained from patients undergoing routine COVID-19 clinical testing at the CLIA and CAP certified laboratories at the Public Health Research Institute (PHRI) and University Hospital, Newark, NJ, were selected for this study. RT-PCR cycle threshold (Ct) values at collection, ranged from a minimum Ct of 12.4 through a maximum Ct of 37.6 and all samples were collected from the months of April through December 2021. Thirty-four of these samples were used for testing in various RT-PCR instruments with all 3 assays. The RT-PCR instruments used were a Roche LightCycler 480 (LC480, Roche, Indianapolis, IN), a Bio-Rad CFX96 (Bio-Rad, Hercules, California), Applied Biosystems™ 7500 (Thermo Fisher Scientific, Waltham, MA), and a Rotor Gene Q (Qiagen, Valencia, CA) located in PHRI laboratories, NJMS genomic laboratory and the UH molecular diagnostics laboratory. These instruments were selected based on the availability and accessibility for this testing. The remaining 40 specimens collected in November and December 2021 were tested only with the LC480 instrument. The distribution of the number of specimens collected over the months and the range of Ct values are shown the supplementary Fig. 1S. RNA was extracted from all the specimens using a QiaAmp viral RNA isolation kit (Qiagen, Valencia, CA) or a QiaSymphony DSP viral RNA extraction kit in a Qiasymphony automated instrument (Qiagen) according to the manufacturers recommendations, and a 5µl volume of this extracted RNA was added to the one-step RT-PCR mix containing the primers and probes (20). Each specimen was run with all 3 assays (SMB-501, SMB-484, and SMB-452) in separate wells. All instruments were programmed with the similar protocol as LC480 as mentioned in the Table 1. The internal control targeting RNAseP was run for all samples. A reference Tm code was established for each SMB assay on all platforms using the WT genomic RNA and corresponding MT SARS-CoV-2 strains. Samples that tested positive either for 501N/484E/452L wildtype or 501Y/484K/484A/452R mutant were confirmed by Sanger sequencing at the Department of Genomic Medicine, Rutgers Biomedical and Health Sciences, Newark using the primer pair: F-5’aggctgcgttatagcttgga3’ and R-5’aaacagttgctggtgcatgt3’ which amplifies a 284bp segment of the S-gene inclusive of the amino acid positions at 452, 484, and 501. Sequencing data were analyzed using Ugene (ver 37) or MegAlign Pro software (DNAStar, ver16).

### Statistical analysis

Standard statistical analyses (average, standard deviation) and graphing were performed using Microsoft excel (ver 2102), GraphPad Prism 8.4.3 for Windows, R version 4.1.1 and ggplot2 package.

## Results

### Limit of detection

The limit of detection (LOD) for the codon 452 (SMB-452 assay) was established with serial log dilutions of the Delta reference strain (NR-5561) RNA in the negative matrix at concentrations ranging from 10^5^ through 0.1 genome equivalents (GE)/reaction in Roche LC480. Each dilution was tested in replicates of four. The data was analyzed and the LOD was established based on the Tm values and Tm peak heights (MPH) from both wild type (WT) and mutant (MT) probes (Fig. 1A). The Tm values for the WT probe (Cy3, λ533-580) and MT probe (Cy5, λ618-660) in the presence of the Delta target were 58.3°C±0.12 and 63.2°C±0.13, respectively defining a mutant detection signature for 452 assay. The overall LOD for the SMB-452 assay, determined as the lowest target concentration where all 4 replicates were positive was found to be 1 GE (Fig. 1A). The LOD for detecting mutations in codons 501 (SMB-501 assay) and 484 (SMB-484 assay) in our previously published test (20) was re-tested with Omicron RNA in 10-fold dilutions ranging from concentrations of 10^5^-1 GE/reaction (Fig. 1B and C) in the presence of COVID-19 negative nasal swab matrix. This testing established the SMB-501 assay LOD as 100 GE/reaction (Fig. 1B) and the SMB-484 assay LOD as 10^3^ GE/reaction (Fig. 1C). Although, 3 out of 4 (75%) at 100 GE/reaction was positive for SMB-484 assay and further work is underway to improve the SMB-484 LOD for Omicron, clinical sensitivity of the assay appears to be excellent (see below).

**Fig. 1.**
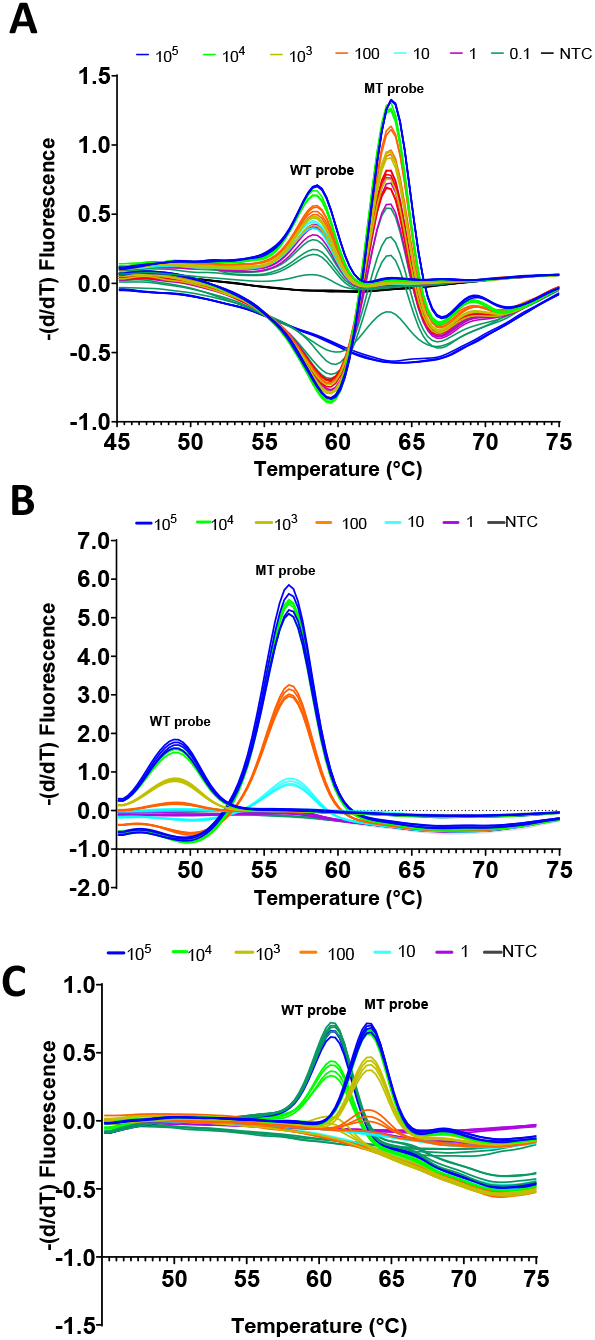
Analytical limit of detection for B.1.617.2 (Delta) RNA tested by SMB-452 assay (A) and B.1.529 (Omicron,) RNA tested by SMB-501 and SMB-484 assays (B and C), in presence of the nasopharyngeal (NP) matrix. The Tm values with wild type specific probe (WT probe) and mutant specific probe (MT probe) shows the signature for the variant strain from the respective SMB assay. RNA tested at different concentrations are shown as number of genomic equivalents (GEs) per 20 µl reaction; NTC-No template control.

### Tm code definition

The Tm values produced by all SMBs against the reference WT and the MT SARS-CoV-2 strains generated in different instruments are listed in Table 2. Tm values can vary slightly between samples and different instrument platforms. The 2 or 3-probe Tm coding approach explained previously (20) provides for a robust sequence identification even in the presence of these Tm fluctuations as shown in Table 2. For example, in the LC480 instrument, both WT and Delta have the same WT 501N allele which results in a mean Tm of 59.7±0.1 for the 501-WT probe and 58.9±0.07 with the 501-MT probe, but Alpha and Beta have a mutant 501Y allele, which results in a mean Tm of 55.7±0.04 for the 501-WT probe and 62.6±0.08°C with the 501-MT probe. Omicron has a highly mutant 501Y allele which results in a mean Tm of 49°C±0.12 for the 501-WT probe and 56.6°C±0.15 for the 501-MT probe. Similarly, Tm codes were established for other codons and both WT and variant alleles as also shown in Table 2. The six-Tm code generated by all three assays was recorded and used as the reference Tm signature for each VOC.

**Table 3.**
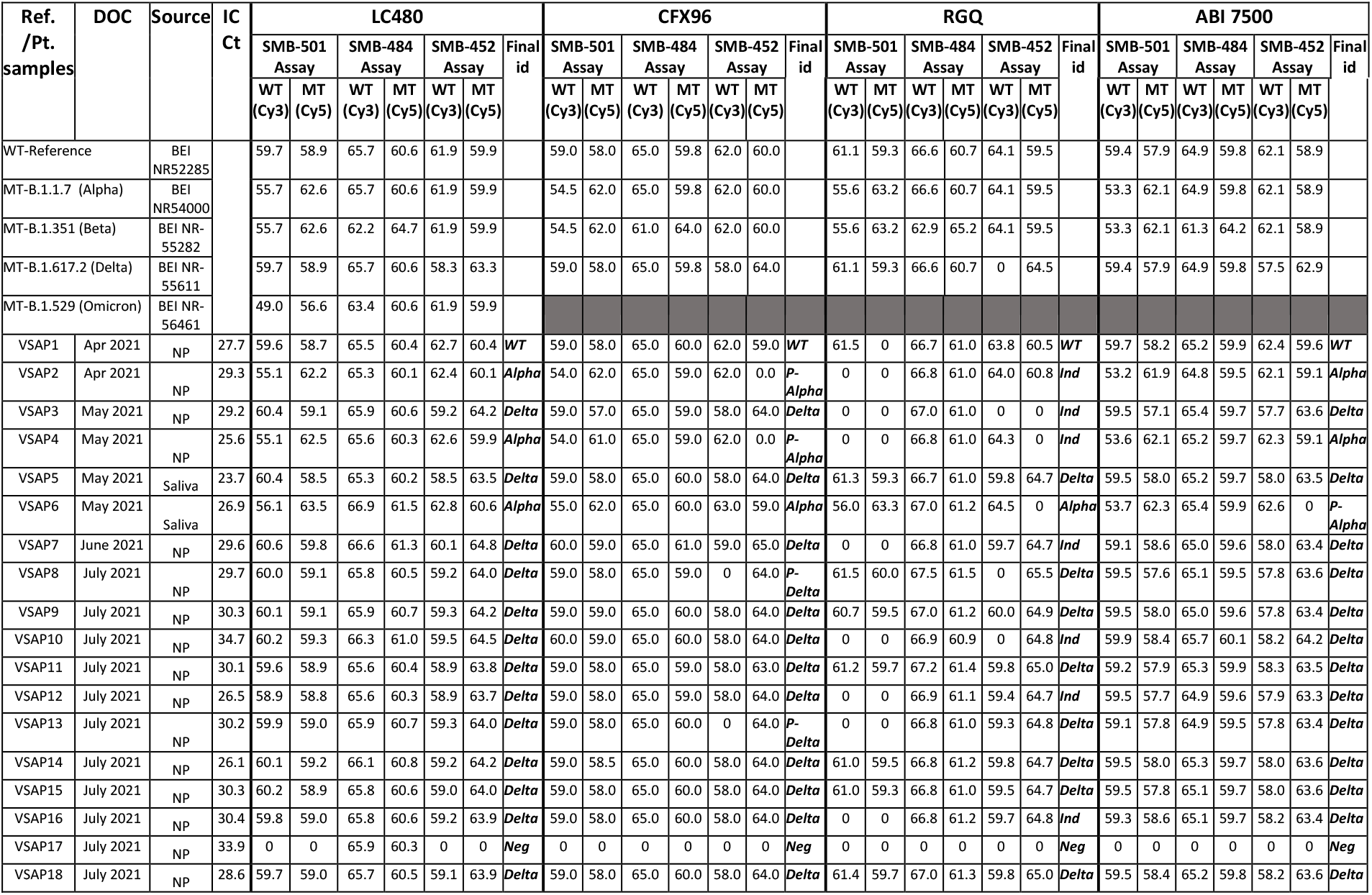
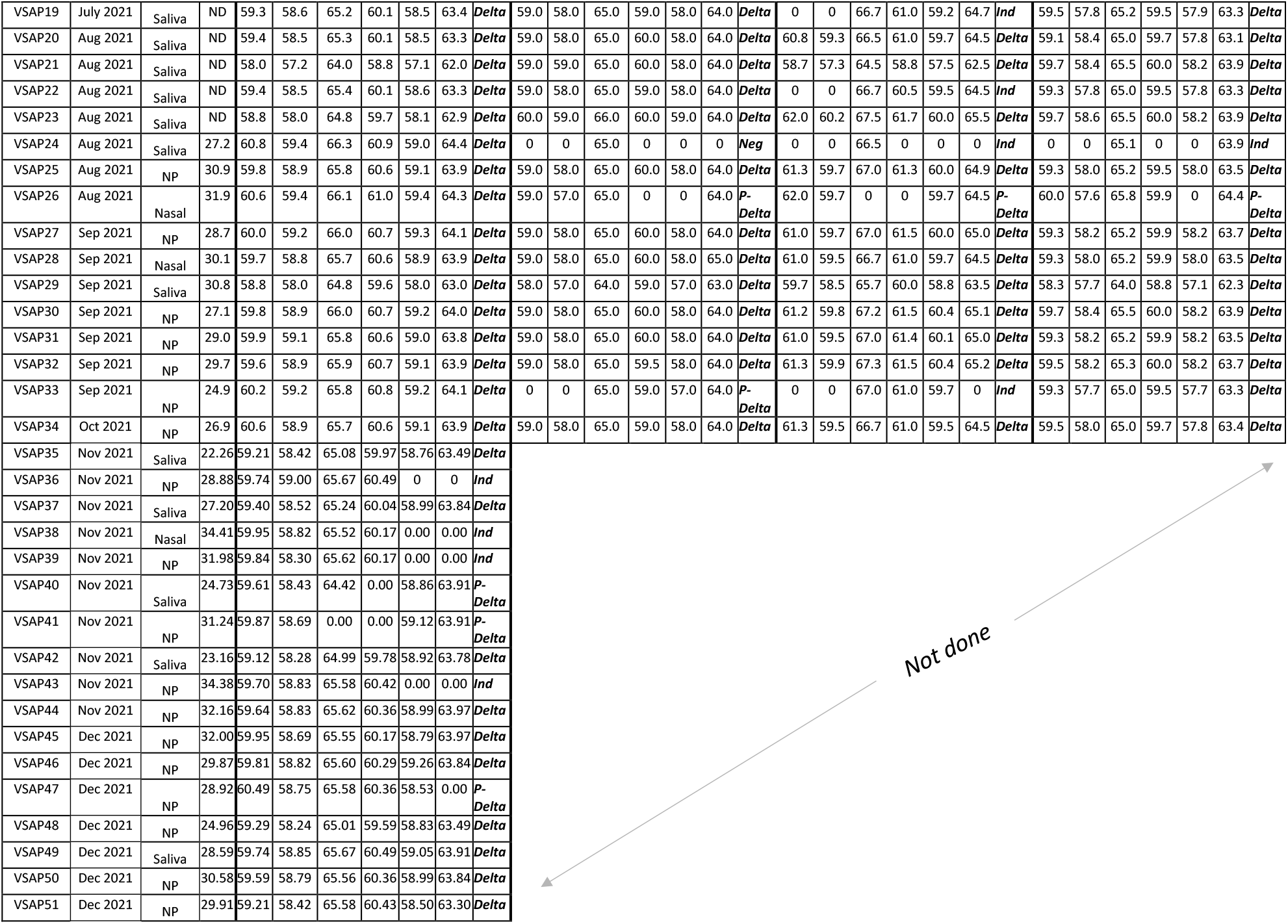
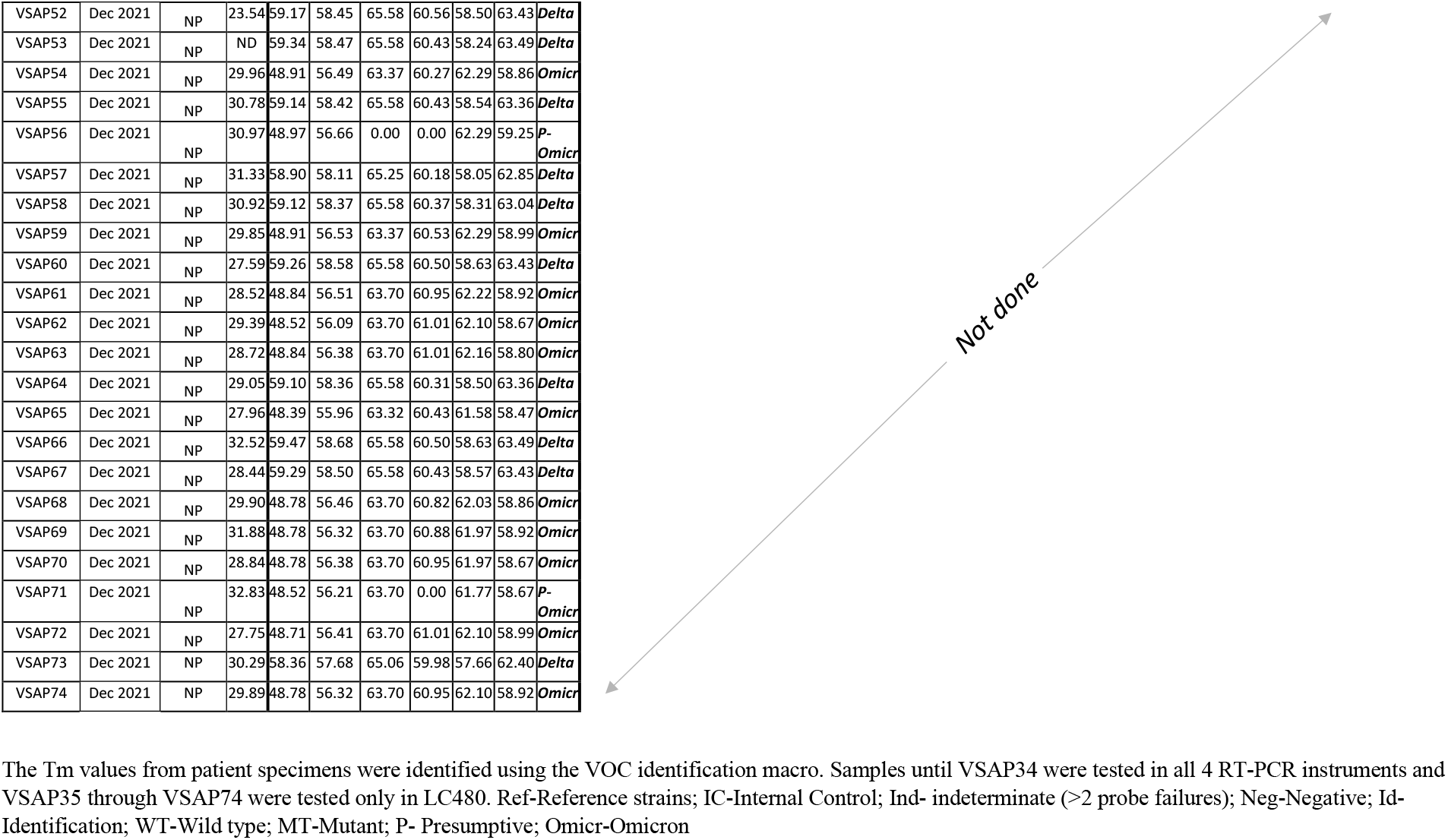
Patient specimen validation in four different RT-PCR instruments with melt curve analysis using 3 SMB assays for detection and differentiation of SARS-Cov-2 variants of concern.

### Testing clinical samples

As shown in Table 2, a total of 74 confirmed COVID-19 positive patient samples were tested out of which 34 were tested with all three defined assays (SMB-501, SMB-484, and SMB-452) on four different RT-PCR instruments with melt analysis capability. The remaining 40 positive samples and 5 confirmed negative samples were tested with the LC480 only, and these results were used for the primary clinical performance analysis. The sample collection timeline and the initial Ct value at collection is shown in supplementary Fig. 1S. We tested the samples collected over 9 months from April to December 2021, and Ct values ranged from 12 to 37.6. All identification for the VOC were made using the Excel analyze tool. The Tm values obtained from all 3 assays, and identification for each patient sample tested are listed in Table 2. The clustering of various mutations and the establishment of a Tm signature to identify specific WT or VOCs using SARS-CoV-2 SMB genotyping test, is demonstrated in Fig. 2. Sanger sequencing of all samples (Table 3) was used to confirm the strain identity of each sample. Compared to Sanger Sequencing, the LC480 correctly identified 49/53 (92%) of the Delta variants as Delta, and correctly identified all 13/13 (100%) Omicron variants as Omicron. Of the four samples not identified as Delta by both the assay and Sanger sequencing, three of the specimens were identified as Delta by the SMB-assay, but failed sequencing, and for of the specimens identified as Delta by sequencing produced indeterminate results by the SMB assay (Table 3). None of the samples were incorrectly identified as WT or incorrectly identified as a different VOC. Thus, excluding samples with indeterminant results, the assay was 100% sensitive and 100% specific. Including the four indeterminate samples reduced assay sensitivity to 94%, without changing the specificity.

**Fig. 2.**
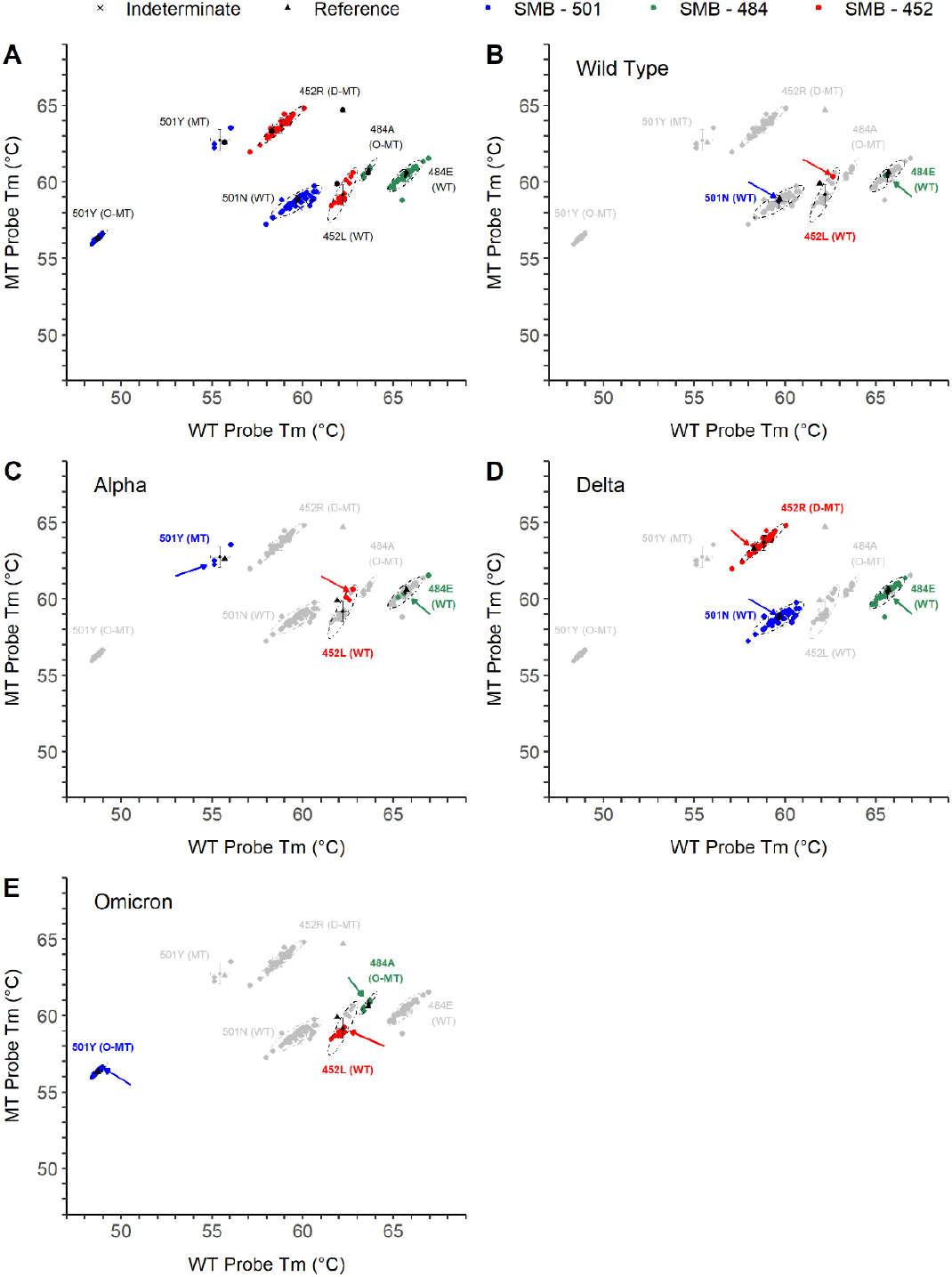
Variant of concern (VOC) detection using the SARS-COV-2 SMB genotyping test. Correlation plots showing grouping and classification of patient specimens (N=74) tested in Roche LC480 instrument using SMB-501 (blue), SMB-484 (Green) and SMB-452 (Red) assays (A). Identification Tm signature specific for the wild type (WT, B); Alpha (C); Delta (D) and Omicron (E) are highlighted and indicated by arrows.

**Table 3.**
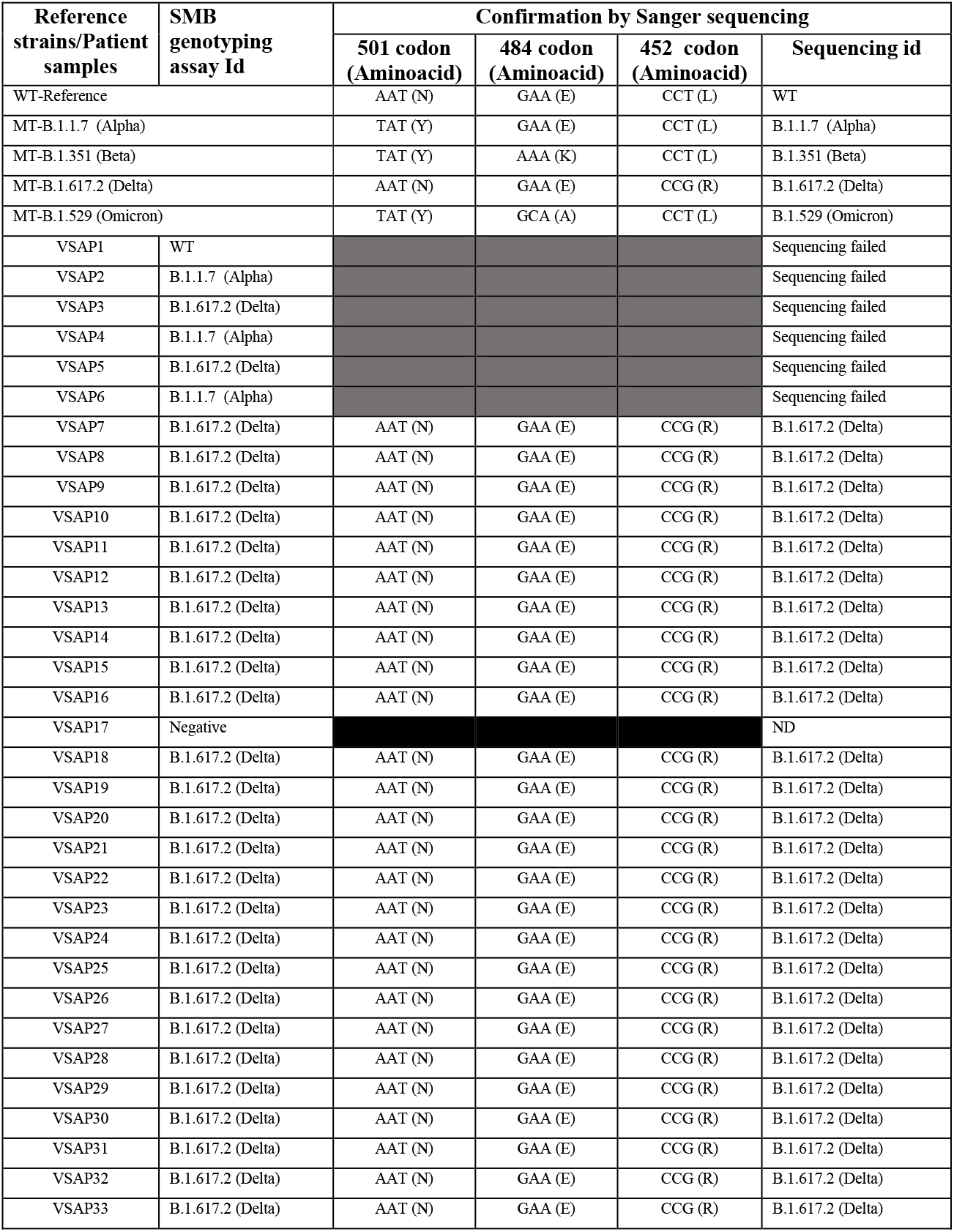
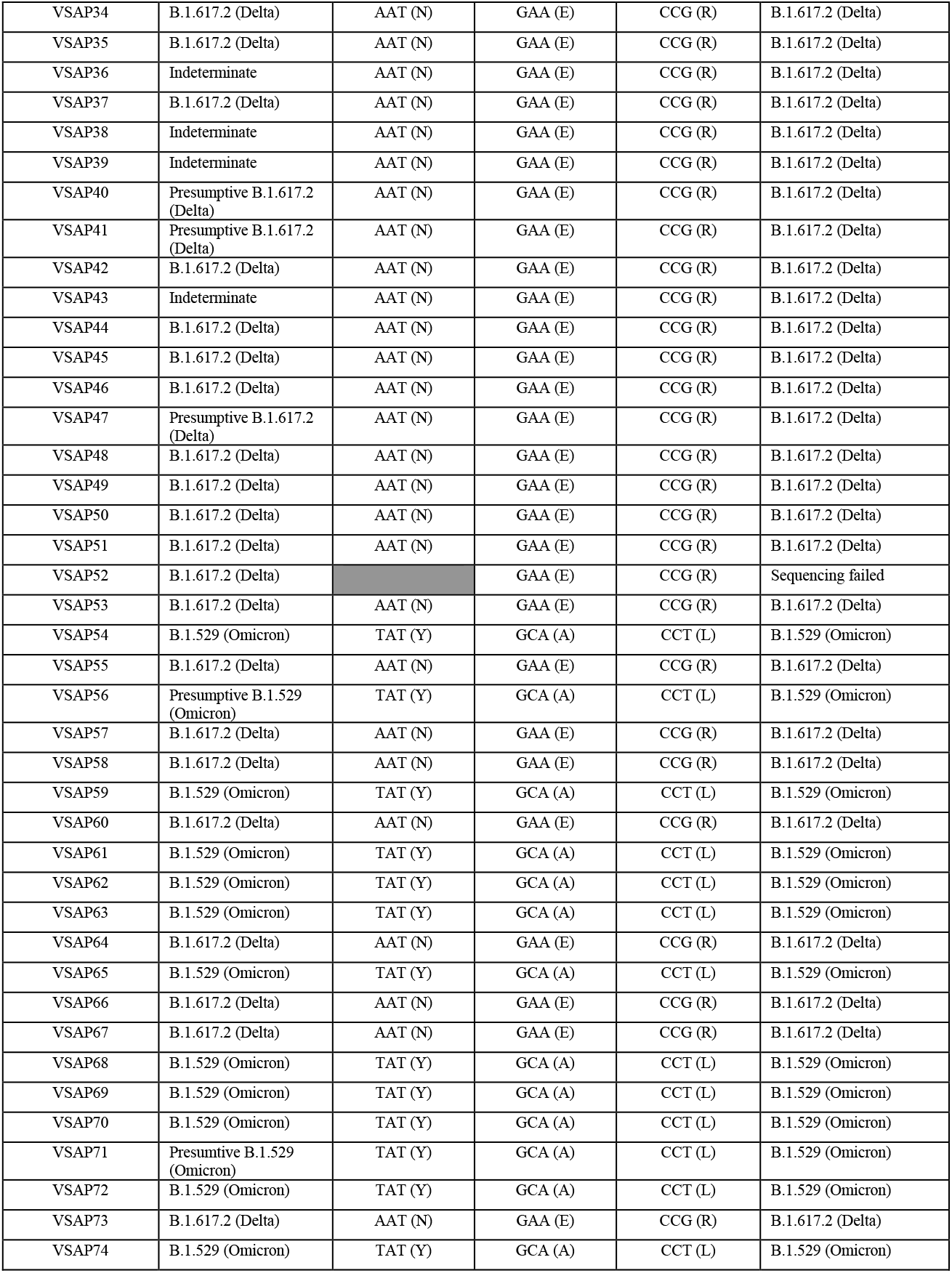
Confirmation of mutations in patient samples by sequencing.

| Reference strains/Patient samples | SMB genotyping assay Id | Confirmation by Sanger sequencing |  |  |  |
| --- | --- | --- | --- | --- | --- |
|  |  | 501 codon (Aminoacid) | 484 codon (Aminoacid) | 452 codon (Aminoacid) | Sequencing id |
| WT-Reference |  | AAT (N) | GAA (E) | CCT (L) | WT |
| MT-B.1.1.7 (Alpha) |  | TAT (Y) | GAA (E) | CCT (L) | B.1.1.7 (Alpha) |
| MT-B.1.351 (Beta) |  | TAT (Y) | AAA (K) | CCT (L) | B.1.351 (Beta) |
| MT-B.1.617.2 (Delta) |  | AAT (N) | GAA (E) | CCG (R) | B.1.617.2 (Delta) |
| MT-B.1.529 (Omicron) |  | TAT (Y) | GCA (A) | CCT (L) | B.1.529 (Omicron) |
| VSAP1 | WT |  |  |  | Sequencing failed |
| VSAP2 | B.1.1.7 (Alpha) |  |  |  | Sequencing failed |
| VSAP3 | B.1.617.2 (Delta) |  |  |  | Sequencing failed |
| VSAP4 | B.1.1.7 (Alpha) |  |  |  | Sequencing failed |
| VSAP5 | B.1.617.2 (Delta) |  |  |  | Sequencing failed |
| VSAP6 | B.1.1.7 (Alpha) |  |  |  | Sequencing failed |
| VSAP7 | B.1.617.2 (Delta) | AAT (N) | GAA (E) | CCG (R) | B.1.617.2 (Delta) |
| VSAP8 | B.1.617.2 (Delta) | AAT (N) | GAA (E) | CCG (R) | B.1.617.2 (Delta) |
| VSAP9 | B.1.617.2 (Delta) | AAT (N) | GAA (E) | CCG (R) | B.1.617.2 (Delta) |
| VSAP10 | B.1.617.2 (Delta) | AAT (N) | GAA (E) | CCG (R) | B.1.617.2 (Delta) |
| VSAP11 | B.1.617.2 (Delta) | AAT (N) | GAA (E) | CCG (R) | B.1.617.2 (Delta) |
| VSAP12 | B.1.617.2 (Delta) | AAT (N) | GAA (E) | CCG (R) | B.1.617.2 (Delta) |
| VSAP13 | B.1.617.2 (Delta) | AAT (N) | GAA (E) | CCG (R) | B.1.617.2 (Delta) |
| VSAP14 | B.1.617.2 (Delta) | AAT (N) | GAA (E) | CCG (R) | B.1.617.2 (Delta) |
| VSAP15 | B.1.617.2 (Delta) | AAT (N) | GAA (E) | CCG (R) | B.1.617.2 (Delta) |
| VSAP16 | B.1.617.2 (Delta) | AAT (N) | GAA (E) | CCG (R) | B.1.617.2 (Delta) |
| VSAP17 | Negative |  |  |  | ND |
| VSAP18 | B.1.617.2 (Delta) | AAT (N) | GAA (E) | CCG (R) | B.1.617.2 (Delta) |
| VSAP19 | B.1.617.2 (Delta) | AAT (N) | GAA (E) | CCG (R) | B.1.617.2 (Delta) |
| VSAP20 | B.1.617.2 (Delta) | AAT (N) | GAA (E) | CCG (R) | B.1.617.2 (Delta) |
| VSAP21 | B.1.617.2 (Delta) | AAT (N) | GAA (E) | CCG (R) | B.1.617.2 (Delta) |
| VSAP22 | B.1.617.2 (Delta) | AAT (N) | GAA (E) | CCG (R) | B.1.617.2 (Delta) |
| VSAP23 | B.1.617.2 (Delta) | AAT (N) | GAA (E) | CCG (R) | B.1.617.2 (Delta) |
| VSAP24 | B.1.617.2 (Delta) | AAT (N) | GAA (E) | CCG (R) | B.1.617.2 (Delta) |
| VSAP25 | B.1.617.2 (Delta) | AAT (N) | GAA (E) | CCG (R) | B.1.617.2 (Delta) |
| VSAP26 | B.1.617.2 (Delta) | AAT (N) | GAA (E) | CCG (R) | B.1.617.2 (Delta) |
| VSAP27 | B.1.617.2 (Delta) | AAT (N) | GAA (E) | CCG (R) | B.1.617.2 (Delta) |
| VSAP28 | B.1.617.2 (Delta) | AAT (N) | GAA (E) | CCG (R) | B.1.617.2 (Delta) |
| VSAP29 | B.1.617.2 (Delta) | AAT (N) | GAA (E) | CCG (R) | B.1.617.2 (Delta) |
| VSAP30 | B.1.617.2 (Delta) | AAT (N) | GAA (E) | CCG (R) | B.1.617.2 (Delta) |
| VSAP31 | B.1.617.2 (Delta) | AAT (N) | GAA (E) | CCG (R) | B.1.617.2 (Delta) |
| VSAP32 | B.1.617.2 (Delta) | AAT (N) | GAA (E) | CCG (R) | B.1.617.2 (Delta) |
| VSAP33 | B.1.617.2 (Delta) | AAT (N) | GAA (E) | CCG (R) | B.1.617.2 (Delta) |
| VSAP34 | B.1.617.2 (Delta) | AAT (N) | GAA (E) | CCG (R) | B.1.617.2 (Delta) |
| VSAP35 | B.1.617.2 (Delta) | AAT (N) | GAA (E) | CCG (R) | B.1.617.2 (Delta) |
| VSAP36 | Indeterminate | AAT (N) | GAA (E) | CCG (R) | B.1.617.2 (Delta) |
| VSAP37 | B.1.617.2 (Delta) | AAT (N) | GAA (E) | CCG (R) | B.1.617.2 (Delta) |
| VSAP38 | Indeterminate | AAT (N) | GAA (E) | CCG (R) | B.1.617.2 (Delta) |
| VSAP39 | Indeterminate | AAT (N) | GAA (E) | CCG (R) | B.1.617.2 (Delta) |
| VSAP40 | Presumptive B.1.617.2 (Delta) | AAT (N) | GAA (E) | CCG (R) | B.1.617.2 (Delta) |
| VSAP41 | Presumptive B.1.617.2 (Delta) | AAT (N) | GAA (E) | CCG (R) | B.1.617.2 (Delta) |
| VSAP42 | B.1.617.2 (Delta) | AAT (N) | GAA (E) | CCG (R) | B.1.617.2 (Delta) |
| VSAP43 | Indeterminate | AAT (N) | GAA (E) | CCG (R) | B.1.617.2 (Delta) |
| VSAP44 | B.1.617.2 (Delta) | AAT (N) | GAA (E) | CCG (R) | B.1.617.2 (Delta) |
| VSAP45 | B.1.617.2 (Delta) | AAT (N) | GAA (E) | CCG (R) | B.1.617.2 (Delta) |
| VSAP46 | B.1.617.2 (Delta) | AAT (N) | GAA (E) | CCG (R) | B.1.617.2 (Delta) |
| VSAP47 | Presumptive B.1.617.2 (Delta) | AAT (N) | GAA (E) | CCG (R) | B.1.617.2 (Delta) |
| VSAP48 | B.1.617.2 (Delta) | AAT (N) | GAA (E) | CCG (R) | B.1.617.2 (Delta) |
| VSAP49 | B.1.617.2 (Delta) | AAT (N) | GAA (E) | CCG (R) | B.1.617.2 (Delta) |
| VSAP50 | B.1.617.2 (Delta) | AAT (N) | GAA (E) | CCG (R) | B.1.617.2 (Delta) |
| VSAP51 | B.1.617.2 (Delta) | AAT (N) | GAA (E) | CCG (R) | B.1.617.2 (Delta) |
| VSAP52 | B.1.617.2 (Delta) |  | GAA (E) | CCG (R) | Sequencing failed |
| VSAP53 | B.1.617.2 (Delta) | AAT (N) | GAA (E) | CCG (R) | B.1.617.2 (Delta) |
| VSAP54 | B.1.529 (Omicron) | TAT (Y) | GCA (A) | CCT (L) | B.1.529 (Omicron) |
| VSAP55 | B.1.617.2 (Delta) | AAT (N) | GAA (E) | CCG (R) | B.1.617.2 (Delta) |
| VSAP56 | Presumptive B.1.529 (Omicron) | TAT (Y) | GCA (A) | CCT (L) | B.1.529 (Omicron) |
| VSAP57 | B.1.617.2 (Delta) | AAT (N) | GAA (E) | CCG (R) | B.1.617.2 (Delta) |
| VSAP58 | B.1.617.2 (Delta) | AAT (N) | GAA (E) | CCG (R) | B.1.617.2 (Delta) |
| VSAP59 | B.1.529 (Omicron) | TAT (Y) | GCA (A) | CCT (L) | B.1.529 (Omicron) |
| VSAP60 | B.1.617.2 (Delta) | AAT (N) | GAA (E) | CCG (R) | B.1.617.2 (Delta) |
| VSAP61 | B.1.529 (Omicron) | TAT (Y) | GCA (A) | CCT (L) | B.1.529 (Omicron) |
| VSAP62 | B.1.529 (Omicron) | TAT (Y) | GCA (A) | CCT (L) | B.1.529 (Omicron) |
| VSAP63 | B.1.529 (Omicron) | TAT (Y) | GCA (A) | CCT (L) | B.1.529 (Omicron) |
| VSAP64 | B.1.617.2 (Delta) | AAT (N) | GAA (E) | CCG (R) | B.1.617.2 (Delta) |
| VSAP65 | B.1.529 (Omicron) | TAT (Y) | GCA (A) | CCT (L) | B.1.529 (Omicron) |
| VSAP66 | B.1.617.2 (Delta) | AAT (N) | GAA (E) | CCG (R) | B.1.617.2 (Delta) |
| VSAP67 | B.1.617.2 (Delta) | AAT (N) | GAA (E) | CCG (R) | B.1.617.2 (Delta) |
| VSAP68 | B.1.529 (Omicron) | TAT (Y) | GCA (A) | CCT (L) | B.1.529 (Omicron) |
| VSAP69 | B.1.529 (Omicron) | TAT (Y) | GCA (A) | CCT (L) | B.1.529 (Omicron) |
| VSAP70 | B.1.529 (Omicron) | TAT (Y) | GCA (A) | CCT (L) | B.1.529 (Omicron) |
| VSAP71 | Presumptive B.1.529 (Omicron) | TAT (Y) | GCA (A) | CCT (L) | B.1.529 (Omicron) |
| VSAP72 | B.1.529 (Omicron) | TAT (Y) | GCA (A) | CCT (L) | B.1.529 (Omicron) |
| VSAP73 | B.1.617.2 (Delta) | AAT (N) | GAA (E) | CCG (R) | B.1.617.2 (Delta) |
| VSAP74 | B.1.529 (Omicron) | TAT (Y) | GCA (A) | CCT (L) | B.1.529 (Omicron) |

Performance of the other three test instruments were compared to the LC480 as the gold standard. The BioRadCFX96 identified 97% (33/34) of the samples correctly and one sample tested negative (Kappa, κ=0.9). The ABI7500 also showed a high agreement with the LC480 (κ=0.9), where 32/34 (94%) of the samples were in agreement, and one sample was indeterminate. However, the Qiagen rotor-geneQ (RGQ) instrument produced relatively high indeterminate rates where a variant type could not be called. The RGQ detected 67.6% of the specimen in agreement with LC480 results (κ=0.39) and 11/34 (32.4%) of the samples were indeterminates.

## Discussion

Although whole genome sequencing is a powerful tool to identify new viral lineages, a simple high throughput screening test that accurately identifies VOC provides many advantages. The current study demonstrates that a modification of our simple and easily adoptable assay which was previously developed to identify Alpha, Beta and Gamma strains, can also detect the rapidly emerging Omicron variant. The assay is sensitive, specific and high throughput and can be performed on most qRT-PCR instruments once reference Tm values are established, unlike almost all commercial assays (31, 32). In our previous publication (20), we hypothesized that mutations at the codons 501 and 484 would be common in other emergent SARS-CoV-2 variants, and mutations at both codons are repeated in Omicron. Although Delta predominantly remained wild type at these codons, mutations at 452 codon similar to the CAL.20C variant observed first in California (33), is considered a key Delta-defining mutation. The mutations on these codons have been shown to be responsible for the increased infectivity, transmission, escape humoral immunity and reduced susceptibility to monoclonal antibody treatments (5, 34) and data is not clear yet on their resistance to antivirals. Assays that detect mutations in these codons will help with surveillance to track the variants and may also help guide targeted therapy.

## Supporting information

GISAID acknowledgment Supplementary Table 1S

## Data Availability

All data produced in the present study are contained in the manuscript and will also be available upon reasonable request to the authors

## Acknowledgements

This study was funded by the National Institute of Allergy and Infectious Diseases of the National Institutes of Health under award number R01 AI131617. S.A.E.M. is partly supported by a grant from the National Cancer Institute, National Institutes of Health R01CA227291. The following reagents was obtained through BEI Resources, NIAID, NIH: SARS-Related Coronavirus 2, Isolate hCoV-19/USA/PHC658/2021 (Lineage B.1.617.2; Delta Variant), NR-55611, contributed by Dr. Richard Webby and Dr. Anami Patel. We thank Dr. Vincent Munster, Rocky Mountain laboratories (NIH) for sharing the Omicron variant RNA. The authors wish to thank all the laboratories that contributed sequence data to the GISAID EpiCoV database. A GISAID acknowledgment table reporting the geographic origin and contributions of genomes analyzed in this study is attached as a supplementary Table 1S.

D.A. receives research support and royalty payments from Cepheid, which sells the Xpert Xpress SARS-CoV-2 and Xpert Xpress SARS-CoV-2/Flu/RSV tests., S.C, is an employee of Cepheid and R.J. is a paid consultant for Cepheid.

---

**Supplementary Fig. S1.**
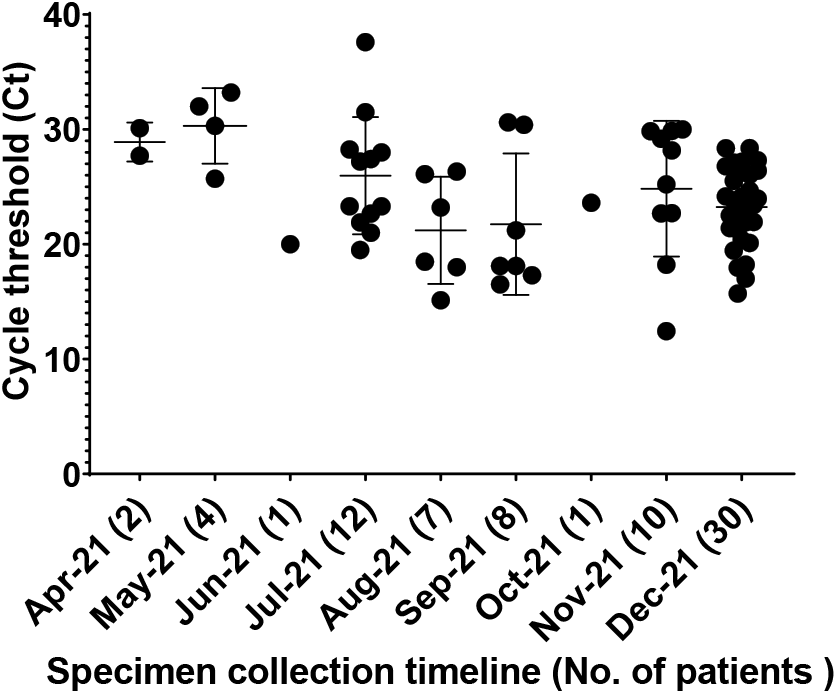
Patient specimen timeline and Cycle threshold (Ct) values at collection.

